# Correlation between Chest CT Severity Scores and the Clinical Parameters of Adult Patients with COVID-19 pneumonia

**DOI:** 10.1101/2020.10.15.20213058

**Authors:** Ghufran Aref Saeed, Waqar Gaba, Asad Shah, Abeer Ahmed Al Helali, Emadullah Raidullah, Ameirah Bader Al Ali, Mohammed Elghazali, Deena Yousef Ahmed, Shaikha Ghanam Al Kaabi, Safaa Almazrouei

**Affiliations:** Department of Radiology, Sheikh Khalifa Medical City, Abu Dhabi, UAE; Department of Internal Medicine, Sheikh Khalifa Medical City, Abu Dhabi, UAE

**Keywords:** COVID-19, CT severity score, High-resolution computed tomography

## Abstract

**Purpose:** Our aim is to correlate the clinical condition of patients with COVID-19 infection with the 25 Point CT severity score by Chang et al (devised for assessment of ARDS in patients with SARS in 2005).

**Material and Methods:** Data of consecutive symptomatic patients who were suspected to have COVID-19 infection and presented to our hospital, was collected from March to April 2020. All patients underwent two consecutive RT-PCR tests and had a non-contrast HRCT scan done at presentation. From the original cohort of 1062 patients, 160 patients were excluded leaving a total number of 902 patients.

**Results:** The mean age was 44.2 ±11.9 years [85.3%males, 14.7%females]. CT severity score found to be positively correlated with lymphopenia, increased serum CRP, d-dimer and ferritin levels (*p* < 0.0001). The oxygen requirements as well as length of hospital stay were increasing with the increase of scan severity.

**Conclusion:** The 25-point CT severity score correlates well with the COVID-19 clinical severity. Our data suggest that chest CT scoring system can aid in predicting COVID-19 disease outcome and significantly correlates with lab tests and oxygen requirements.

## Introduction

Covid-19 is an infection that has widely and rapidly spread all over the world and became a pandemic with significant impacts upon the sociopolitical milieu and healthcare delivery systems (1). The clinical presentations vary from asymptomatic carriers to patients requiring assisted ventilatory support and ICU admissions with increased mortality made it an unusual and unprecedented challenge (2, 3). The nasopharyngeal swab RT-PCR test has been the diagnostic test used as the standard of reference for disease confirmation (4). Although the test is a powerful tool, however there is a small but significant proportion of false negative results reported (5).

A non-contrast High resolution CT chest imaging plays a pivotal and essential role in the early disease detection, particularly in patients with false negative RT-PCR results, as well as in managing and monitoring the course of disease (6). Moreover, the disease severity can be ascertained from the imaging findings, significantly supporting the clinicians in their clinical judgment and ensuring effective and timely management (7). Prognosis can also be affected by the severity of the disease in the critically ill patients allowing appropriate selection of early involvement of the intensive care (8, 9).

Multiple studies have explored the pulmonary involvement on the chest CT images using both visual and software quantitative assessments (10, 11). To our knowledge, ours is the first comprehensive study to describe the correlation of chest CT severity scores and the clinical picture of patients with COVID-19 disease in the Gulf and Arab region. Our study correlates the CT severity score with the clinical severity of the patients who were confirmed to have COVID-19 disease using the 25-point visual quantitative assessment.

## Methods

### Data collection

Ethical approval was obtained from Institutional Review Board (IRB) and Department of health (DOH), Abu Dhabi, United Arab Emirates (UAE). The informed consent was waved off as per the Ethics committee. We collected clinical and laboratory data for analysis, derived from an electronic medical record system, from March to April 2020 of patients who were suspected to have COVID-19 infection and underwent a chest HRCT scan. The results for the chest HRCT images were collected and evaluated using the Picture Archiving and Communication Systems (PACS).

### HRCT inspection

All initial chest HRCT scans were performed on the day of patients’ presentation using a VCT GE 64 scanner. Patients were placed in a supine position with single breath hold. Scanning parameters were: scan direction (craniocaudally), tube voltage (120KV), tube current (100-600 mA)-smart mA dose modulation, slice collimation (64 × 0.625 mm), width (0.625 × 0.625 mm), pitch (1), rotation time (0.5 s), scan length (60.00 – I300.00 s).

### HRCT image analysis

Two radiologists with more than 8 years of experience evaluated the images to determine the disease severity score in each patient. The scans were first assessed whether negative or positive for typical findings of COVID19 pneumonia (bilateral, multilobe, posterior peripheral ground glass opacities) as defined by the ***RSNA Consensus statement*** (7, 12). Severity then was assessed using the following scoring system which depends on the visual assessment of each lobe involved (13, 14, 15), (Figure1):

**Figure 1:**
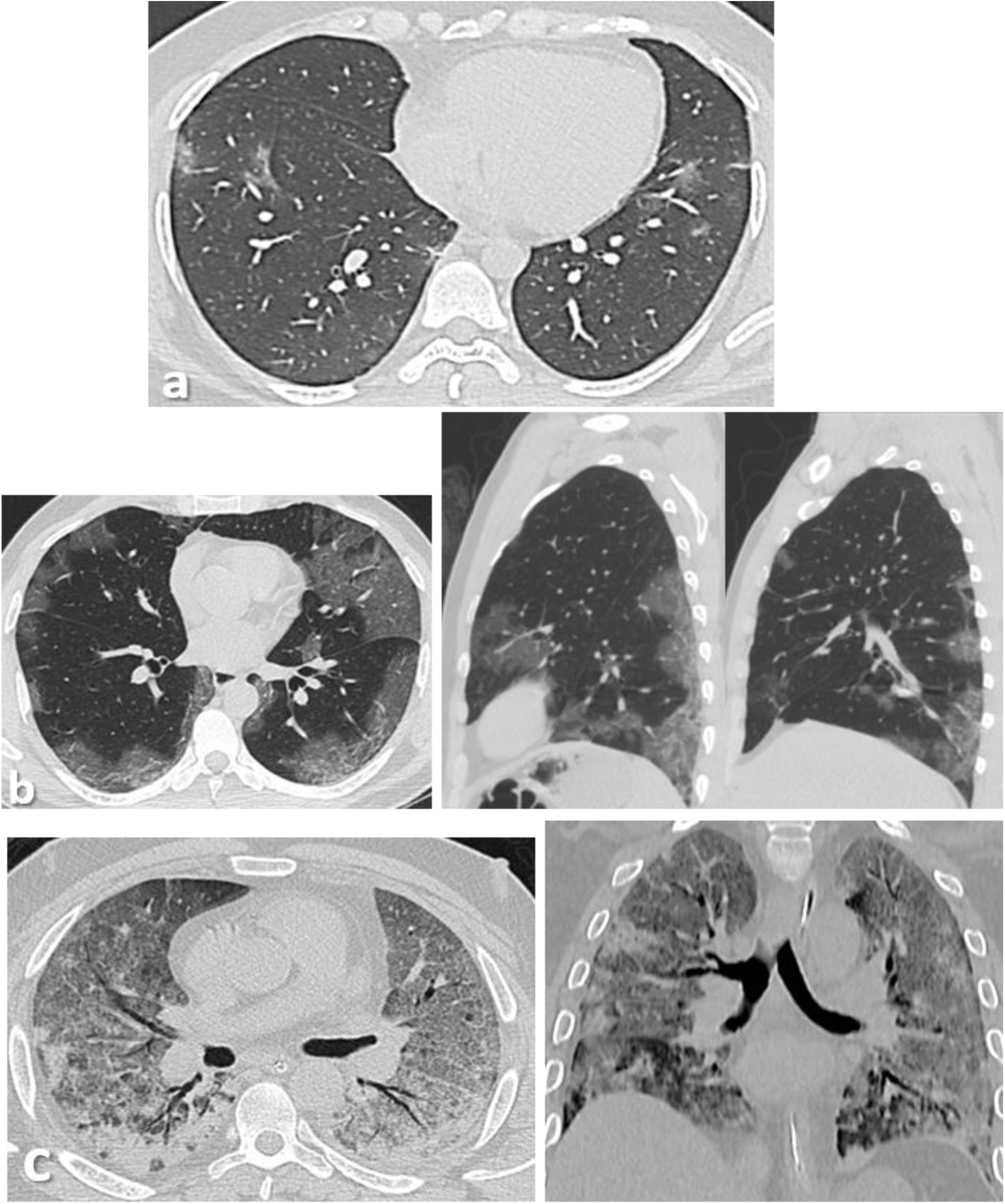
(a) Axial thin-sections of unenhanced CT chest scan show mild GGO involving bilateral peripheral lower lobes. (b) Axial and sagittal sections show bilateral peripheral multilobe GGO of moderate disease severity. (c) Axial and coronal sections show diffuse crazy-paving pattern with areas of peripheral consolidations indicating severe disease.

| Percentage of lobar involvement | Score |
| --- | --- |
| 5% or less | 1 |
| 5%-25% | 2 |
| 26%-49% | 3 |
| 50%-75% | 4 |
| > 75%. | 5 |

The sum of the lobar scores indicates the overall severity:

| Total score (numerical) | Severity (category) |
| --- | --- |
| 7 or less | Mild |
| 8-17 | Moderate |
| 18 or more | Severe |

### Statistical analysis

The analysis was performed using SPSS 21.0. Descriptive statistics of patients’ demographics, clinical, and laboratory results were reported as numbers and relative frequencies. Frequencies of CT scores were calculated and compared with other clinical variables. Pearson correlation coefficient test was used for correlations and p-value less than 0.05 defined statistical significance.

## Results

### Baseline information

Our population included 1,062 consecutive patients who were suspected to have COVID-19 infection. Infection with SARS-CoV-2 was confirmed from a nasopharyngeal swab using the U-TOP COVID-19 Detection Kit (Seasun Biometerials Inc., Daejeon, Korea), which is a reverse transcriptase-polymerase chain reaction (RT-PCR) test that has received Emergency Use Authorization (EUA) from the US Food and Drug Administration (FDA). RT-PCR testing was performed using Clinical Laboratory Improvement Amendments (CLIA) diagnostic standards according to current testing guidelines (16). All patients underwent two consecutive RT-PCR tests and had a HRCT scan done.

160 patients were excluded from the study as per the following *Exclusion Criteria*: patients less than 18 years old, patients with negative RT-PCT results, discharge to another facility leading to lost follow up, suboptimal HRCT scan due to significant motion artefacts or CT with atypical findings for COVID19 pneumonia.

Eventually, 902 patients were included with the following information been collected: age, gender, presence of comorbidities/risk factors, laboratory tests including lymphocyte count, CRP, d-dimer and ferritin levels, maximum O2 requirement, need for intubation, ICU admission, length of hospital stay (LOS) and final clinical outcome (alive, expired, or still intubated at the time of study).

The mean age was 44.2 ±11.9 years [range 19-87 years, 769 males (85.3%), 133 females (14.7%)]. The age was further classified into 6 groups: (<30, 30-39, 40-49, 50-56, 60-69, >70 years).

The risk factors considered included hypertension, diabetes mellitus, asthma, COPD, coronary artery disease, chronic kidney disease. Risk factors were found in 399/902 patients (44.2%) [one risk factor n=206 (22.8%), two risk factors n=114 (12.6%), three or more risk factors n=79 (8.8%)].

Laboratory results showed lymphopenia (normal value 1.5-4 × 109/L) in 203 patients (22.5%), elevated CRP (50 mg/L) in 236 patients (40.3%), high d-dimer (>1 mcg/mL) in 147 (16.2%), and elevated ferritin level (>600 ng/mL) in 301 (33.3%).

Out of the 902 patients, 646 patients (71.6%) didn’t require any oxygen support. The remaining 256 patients required oxygen supplement as follows: 126 patients (14%) required nasal canula, 32 patients (3.5%) required facemask, 15 patients (1.7%) required non-breather mask, 21 patients (2.3%) required a bilevel positive airway pressure (BiPAP) or a high flow nasal cannula (HFNC), 62 patients (6.9%) required intubation, out of which 32 patients (51.6%) were eventually extubated. ICU admission was required in 202 (22.4%) out of the 902 patients with a male predominance (182/202; 90.1%). The commonest age group was that between 40-49 years old (53/202; 26.2%).

Regarding hospital stay, 631 patients (70%) stayed in the hospital for 5 days or less, 127 (14.1%) for 6-11 days, 50 (5.5%) for 11-15 days and 94 (10.4%) for more than 16 days.

In terms of clinical outcome, 863 patients (95.7%) were found alive, 33 (3.7%) expired in hospital and 6 (0.7%) were still intubated at the time of the study.

### Correlation between CT severity and clinical parameters

#### Age and Gender

Our results showed significant correlation (p<0.05) between CT severity score and the male gender, raised inflammatory markers, maximum O2 requirement, length of hospital stay (LOS), need for intubation, and clinical outcome.

The HRCT scans were negative in 203/202 (22.5%) of patients (75.9% males, 24.1% females). Mild disease was seen in 329/209 (36.5%) patients (84.5% males, 15.5% females); moderate in 309/902 (34.3%) (90.6 males, 9.4% females); and severe in 61/209 (6.8%) patients (93.4% males, 6.6% females). A follow up scan was done for 73/902 patients (8.1%).

The negative and mild scans were mainly seen in the (30-39 years) age group, (n=62, 30%) and (n=96, 29.2%), respectively; and least in >70 years old (n=6, 3%) and (n=4, 1.2%), respectively. Moderate disease severity was mainly seen in the (40-49 years) age group (n=98, 31.7%) and least in >70 years old (n=4, 1.3%). Severe disease was mainly seen in the (50-59 years) age group (n=21, 34.4%) and least in (18-29 years), (n=0, 0%) (Figure2). The highest percentage of patients with 3 or more risk factors was seen in the severe group (11.5%).

**Figure 2:**
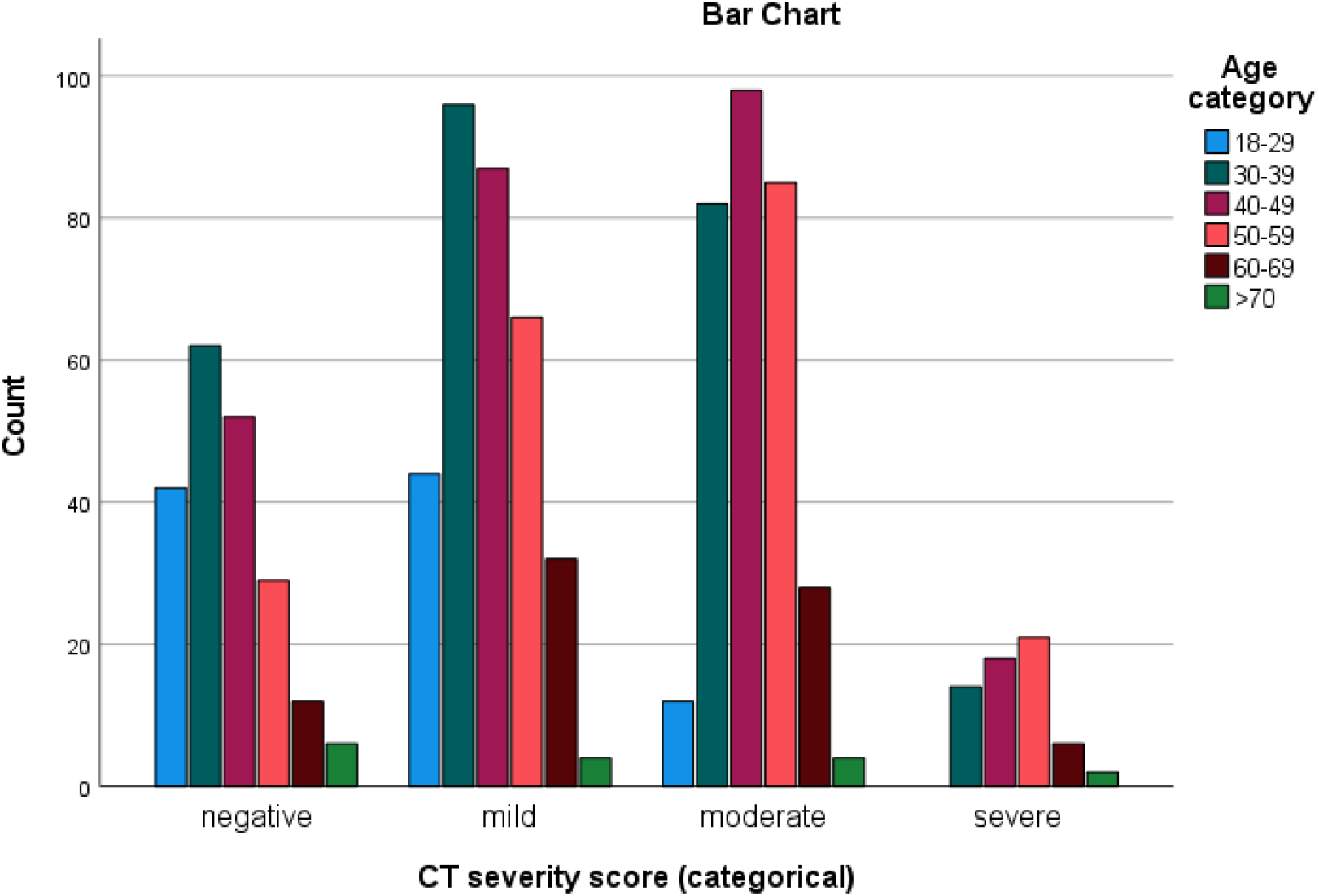
Prevalence of age categories for each CT severity score.

#### Laboratory results

Lymphopenia was detected in 17 patients (8.4%) of the negative group; 55 patients (13.7%) of the mild group; 110 patients (35.6%) of the moderate group; and 31 patients (50.8%) of the severe group. When compared with disease phase, lymphopenia found to be more significant among patients with more severe scans (p < 0.0001).

CRP results were available for 897/902 patients. In patients with negative CT scan, CRP levels were <50mg/L in 193 patients (96%), and in 308 patients (94.2%) with mild scan results. In patients with moderate CT scan, CRP levels were <50mg/L in 151 patients (49%), 50-100mg/L in 93 patients (30.2%), and >100mg/L in 64 patients (20.8%). In patients with severe CT scan, CRP levels were <50mg/L in 9 patients (14.8%), 50-100mg/L in 13 patients (21.3%), and >100mg/L in 39 patients (63.9%). These findings were found to have statistically significant correlation with the CT severity score (p < 0.0001, r = 0.556).

Moreover, the d-dimer level was assessed in 890/902 patients. Most patients with negative CT studies and mild CT severity scores had a normal d-dimer level; 182 patients (94.3%), and 304 patients (93%), respectively. In moderate category, the level was <1mcg/mL in 239 patients (77.3%); 1-3mcg/mL in 63 patients (20.4%), and >3mcg/mL in 7 patients (2.3%). In the severe category, the level was <1mcg/mL in 18 patients (29.5%); 1-3mcg/mL in 28 patients (45.9%), and >3mcg/mL in 15 patients (24.6%).

Ferritin level was assessed in (888/902) patients. In patients with negative and mild CT scan findings, ferritin level was <600ng/mL in 176 patients (91.7%) and 267 patients (81.4%), respectively. In patients with moderate CT scans, ferritin level was <600ng/mL in 131 patients (42.7%); 600-1200ng/mL in 99 patients (32.2%); 1200-2400ng/mL in 60 patients (19.5%), and >2400ng/mL in 17 (5.5%). In patients with severe CT scan, ferritin level was <600ng/mL in 13 patients (21.3%); 600-1200ng/mL in 18 patients (29.5%); 1200-2400ng/mL in 18 patients (29.5%), and >2400ng/mL in 12 (19.7%).

#### Oxygen requirement and outcome

In relation to maximum oxygen requirement, 196/203 (96.6%) of patients with negative scan didn’t require any oxygen support; five patients (2.5%) required nasal cannula; one patient (0.5%) required BiPAP/HFNC and one (0.5%) required intubation eventually.

In the mild scan category, 293/329 (89.1%) patients didn’t require any oxygen support; 24 (7.3%) required nasal cannula; four (1.2%) required facemask; one (0.3%) required non-breather mask; one (0.3%) required BiPAP/HFNC and six (1.8%) required intubation.

154/309 (49.8%) of patients with moderate scan findings didn’t require any oxygen support; 89 (28.8%) required nasal cannula; 20 (6.5%) required facemask; 9 (2.9%) required non-breather mask; 15 (4.9%) required BiPAP/HFNC and 22 (7.1%) required intubation.

Only two patients (3.3%) with severe scan results didn’t require oxygen support; 8 (13.1%) required nasal cannula; 8 (13.1%) required facemask; 5 (8.2%) required non-breather mask; 4 (6.6%) required BiPAP/HFNC and 34/61 (55.7%) required intubation (Table 1).

**Table 1:**
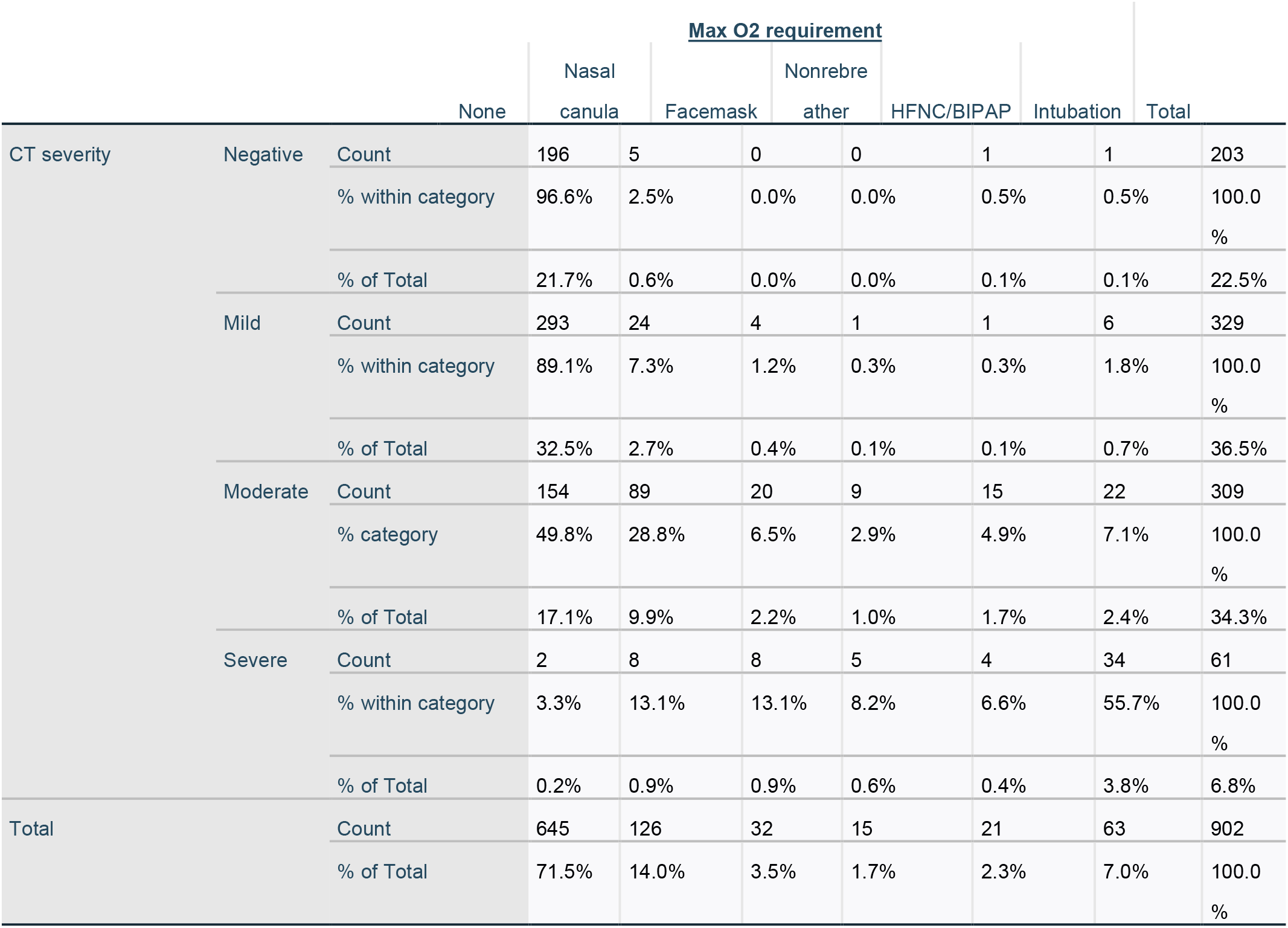
Maximum oxygen requirement in each CT severity category.

The oxygen requirement and CT severity scores were found to have statistically significant correlation (p < 0.0001, r = 0.529).

In terms of length of hospital stay, 190/902 patients (21.1%) with negative scans; 276 patients (30.6%) with mild scans; 160 (17.7%) patients with moderate; and 5 patients (0.6%) with severe findings were either discharged same day or required hospital admission for <5 days.

Six patients (0.7%) stayed in the hospital for 6-10 days from the negative group; 31 (3.4%) from mild; 79 (8.8%) from moderate, and 11 (1.2%) from severe groups.

Four patients (0.4%) stayed in the hospital for 11-15 days from the negative group; 8 (0.9%) from mild; 28 (3.1%) from moderate, and 10 (1.1%) from severe groups.

Three patients (0.3%) stayed in the hospital for >16 days from the negative group; 14 (1.6 %) from mild; 42 (4.7%) from moderate, and 35 (3.9%) from severe groups.

ICU admission was required in 202/902 (22.4%) patients. 23/902 (2.5%) patients with initial negative scan required ICU admission because of subsequent deterioration; 72 (8%) had mild CT scan findings; 69 (7.6%) had moderate; and 38 (4.2%) with severe CT findings (Figure3).

**Figure 3:**
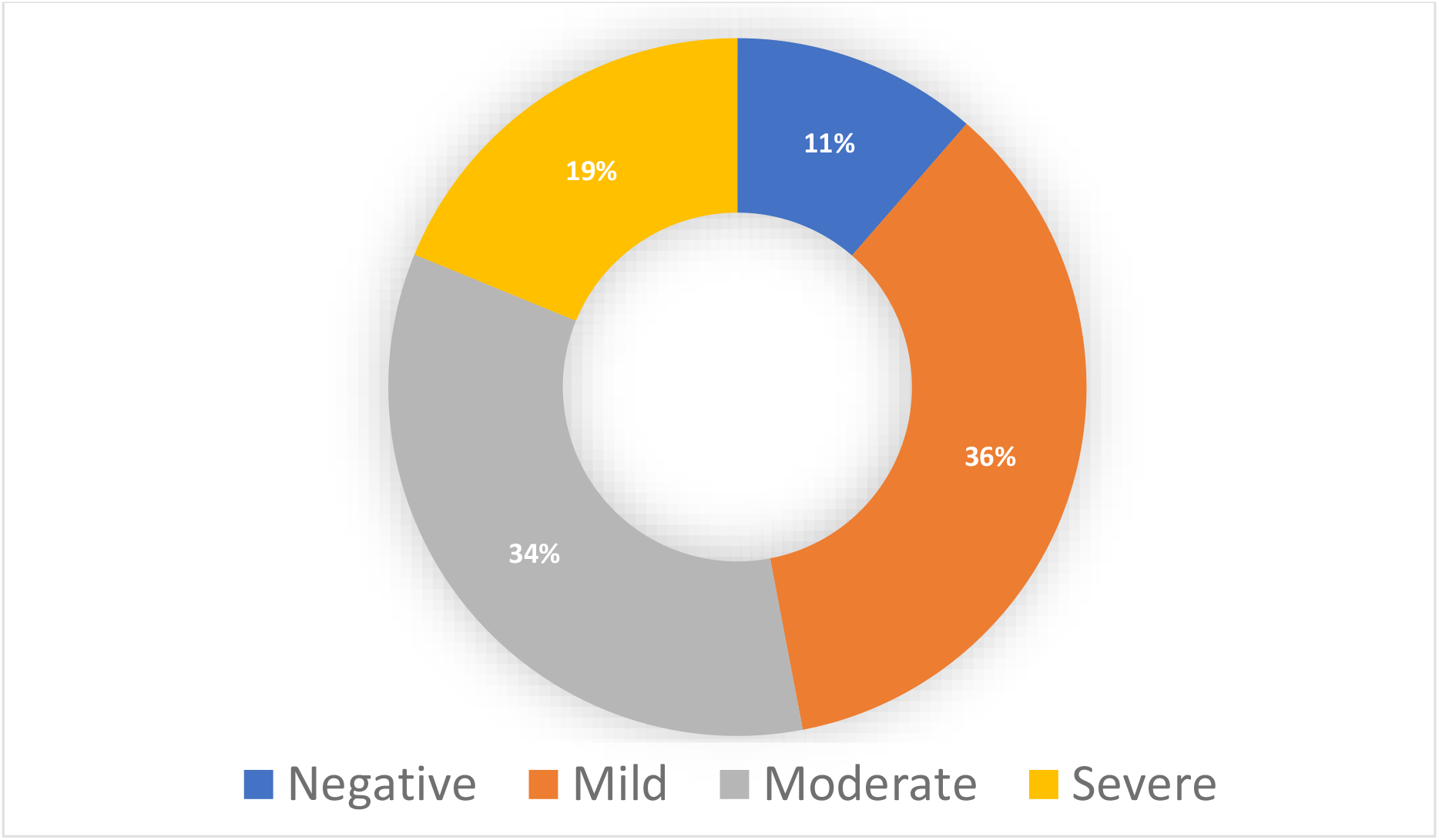
Percentage of patients admitted to the ICU in each CT severity category.

Follow up CT scans (n=73) were done between 7-45 days from initial ones. 18 scans were done for patients who were in the ICU. Four scans (22.2%) showed improvement; 8 (44.4%) showed worsening of the findings and the rest were relatively stable. 55 scans were done for patients who were not admitted to the ICU at that time. Sixteen scans (29.1%) showed improvement; 30 (54.5%) showed worsening and the rest showed stable findings.

The best outcomes were associated with negative and milder CT findings, while death rate was increased among those with more severe scan results. Amongst the negative scan group, 201 patients (99%) are alive, 1 (0.5%) was expired, and 1 (0.5%) is still intubated. Among the mild ones; 325 (98.8%) patients are alive, 3 (0.9%) were expired and one (0.3%) is still intubated. In the moderate scan group; 292 (94.5%) patients are alive, 15 (4.9%) were expired and two (0.6%) are still intubated. In the severe scan group; 44 (72.1%) patients are alive, 14 (23%) were expired and 3 (4.9%) were still intubated at the time of study.

33 patients had in hospital death, with ages ranging between 33-76 years (Mean 55.2 years). 15/33 patients (45.5%) who were expired were between 50-59 years old; 30.3% between 60-69; 15.2 (40-49); 6% (30-39) and 3% >70.

## Discussion

The WHO advised the use of chest imaging as part of diagnostic workup of COVID-19 disease whenever RT-PCR testing is not available; in case of delayed test results or when there is a clinical suspicion of COVID-19 with initial negative RT-PCR testing. Clinicians should work hand in hand with the radiologists in order to make the proper choice of imaging modality (17).

CT scan can be a useful tool in evaluating the individual disease burden (18). The quantitative severity can be assessed using a visual method (as in our study) or a software that determines the percentage of affected lung volumes using the deep learning algorithms (10, 19, 20).

In our study, and due to unavailability of the software, we used the visual assessment of each of the 5 lung lobes. The severities were further classified based on the total cumulative severity score.

Our population revealed a relatively young age (mean 44.2 years) with male predilection. This can be explained by the particular characteristics of the population in the UAE, with prevalence of young male immigrant workers (21).

Severe disease was mostly seen in males (93.4%). Studies suggest that such distribution can be attributed to many factors like disparity in behavior and the possible protective effect of estrogen (22). The most severe disease as well as the highest mortality rates were found in the (50-59 years) age group. This can be affected by different factors like the stage of the pandemic when the study was carried, presence of patients ‘comorbidities, maturity and preparation of the healthcare system, and existence of elderly nursing homes services where disease can spread faster (23).

A number of existing literature suggested that the presence of risk factors, particularly hypertension, diabetes, lung and coronary artery diseases, carries a poor prognosis, with even worse outcome when multiple risk factors are present (24,25). Although in our study we didn’t find a statically significant correlation between the presence of risk factors and CT severity scores, there was however a significant correlation (p< 0.0001) between the ICU admission and presence of risk factors (Figure 4).

**Figure 4:**
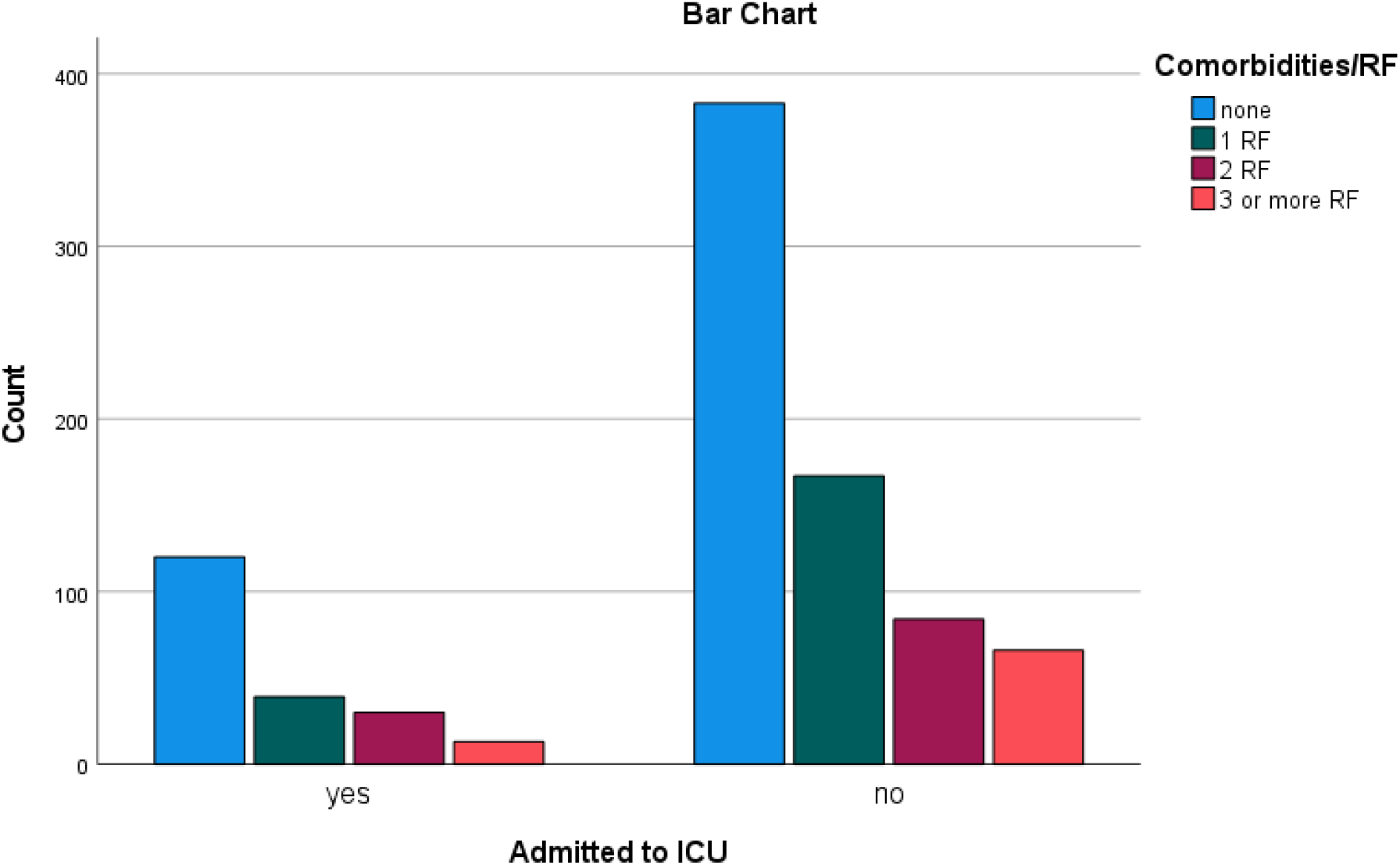
Number of patients admitted to ICU in relation to risk factors.

Lymphopenia correlated well with the increasing CT severity score. The presence of lymphopenia can be related to the inflammatory cytokine storm. Decreased T cell counts, particularly CD8+, has been observed in severe cases (26).

Furthermore, our results showed that the serum CRP level had significant correlation with CT severity. Studies have also suggested that early treatment at early disease stage can be considered using CRP as a predictive marker for likelihood of disease progression (27). Similarly, serum ferritin is a vital mediator of immune dysregulation and its level was closely linked to the severity of the disease (28). D-dimer likewise, can be used as a prognostic indicator, where higher levels are seen in more critical conditions. However, there is lack of evidence regarding the causal effect. It is not yet clear whether this increase is related to the direct effect of the virus or the systemic inflammatory response (29, 30).

As expected, and found in our data, oxygen requirements increase with the increasing in CT severity. The progressive increase in oxygen requirement can be due to the direct damage of the lung by the virus causing inflammatory changes of alveolar wall that limit oxygen exchange, leading to acute respiratory distress, pulmonary fibrosis and eventually death. Moreover, significant pulmonary thromboembolic effects were also found on autopsies from patients who died from COVID-19 disease (31, 32, 33).

Concerning the length of hospital stay for patients with COVID-19 disease, a systematic review done by Rees *et al*. have suggested that LOS varies depending on multiple factors such as: admission and discharge criteria, bed demand and availability, as well as different timing within the pandemic (34). Death rate in our cohort was significantly increased among patients with severe CT findings, as noted in other studies (35).

There are several limitations in this study. First, the need for a larger multicenter cohort to increase the accuracy of the findings. Second, the fact that the assessment of disease severity on CT scans can be subjective. This was reduced by involving two experienced readers to reach a consensus. Finally, the other factors that might contribute to the disease outcome such as lifestyle, as well as relying on self-reporting/ underreporting of the comorbidities should be considered.

In conclusion, CT scans can have a pivotal role in assisting physicians in the management plan as well as work as an indicator for disease severity and possible outcome. CT severity score is positively correlated with inflammatory lab markers, length of hospital stays and oxygen requirement in patients with COVID-19 infection. More studies from different regions would enhance the accuracy of information regarding this novel disease.

## Data Availability

Data available upon request

## Abbreviations

COVID-19: coronavirus disease
CRP: C-reactive protein
CT: computed tomography
GGO: ground-glass opacity
HRCT: high-resolution computed tomography
PACS: picture archiving and communication systems
RT-PCR: reverse transcription polymerase chain reaction
WHO: World Health Organization

## Declaration of Competing Interest

No conflict of interest needs to be disclosed.

## Funding sources

This research did not receive any specific grant from funding agencies in the public, commercial, or not-for-profit sectors.

## Acknowledgments

The authors would like to thank Mr. George Roy, Senior CT radiographer for providing the technical information and Mrs. Maisoun Al Ali, for assisting in the analysis. We would also like to acknowledge the radiological medical and technical team of the Radiology Department of Sheikh Khalifa Medical City-Abu Dhabi.

## Notes

### Competing Interest Statement

The authors have declared no competing interest.

### Clinical Trial

This is a retrospective study

### Author Declarations

Institutional Review Board (IRB) and Department of health (DOH), Abu Dhabi, United Arab Emirates (UAE)

